# Intensity of sample processing methods impacts wastewater SARS-CoV-2 whole genome amplicon sequencing outcomes

**DOI:** 10.1101/2022.09.22.22280217

**Authors:** Shuchen Feng, Sarah M. Owens, Abhilasha Shrestha, Rachel Poretsky, Erica M. Hartmann, George Wells

## Abstract

Wastewater SARS-CoV-2 surveillance has been deployed since the beginning of the COVID-19 pandemic to monitor dynamics in virus burden in local communities. Genomic surveillance of SARS-CoV-2 in wastewater, particularly the efforts for whole genome sequencing for variant tracking or identification, are comparatively challenging due to low target concentration, complex microbial and chemical background, and lack of robust nucleic acid recovery experimental procedures. The intrinsic sample limitations are inherent to wastewater. In this study, we evaluated impacts from sample types, certain sample intrinsic features, and processing and sequencing methods on sequencing outcomes with a specific focus on the breadth of genome coverage. We collected 184 composite and grab wastewater samples from the Chicago area between March to October 2021 for SARS-CoV-2 quantification and genomic surveillance. Samples were processed using a mixture of processing methods reflecting different homogenization intensities (HA+Zymo beads, HA+glass beads, and Nanotrap), and were sequenced using two sequencing library preparation kits (the Illumina COVIDseq kit and the QIAseq DIRECT kit). A synthetic SARS-CoV-2 RNA experiment was performed to validate the potential impacts of processing methods on sequencing outcomes. Our findings suggested that 1) wastewater SARS-CoV-2 whole genome sequencing outcomes were associated with sample types and processing methods 2) in less intensive method processed samples (HA+glass beads), higher genome breadth of coverage in sequencing (over 80%) was associated with N1 concentration > 10^5^ cp/L, while in intensive method (HA+Zymo beads), qPCR results were inconsistent with sequencing outcomes, and 3) sample processing methods and sequencing kits, rather than the extraction methods or intrinsic features of wastewater samples, played important roles in wastewater SARS-CoV-2 amplicon sequencing. Overall, extra attention should be paid to wastewater sample processing (e.g., concentration and homogenization) for sufficient, good quality RNA yield for downstream sequencing.

## Introduction

Since the beginning of the COVID-19 pandemic, wastewater-based epidemiology (WBE) has been applied for the surveillance for SARS-CoV-2 and its variants community-wide^1–4^. In September 2020, wastewater surveillance was officially established at the national level in the United States to support the COVID-19 pandemic response^5^. Earlier wastewater surveillance studies mostly focused on tracking the change of viral concentration in the sewersheds by applying reverse-transcription quantitative polymerase chain reaction (RT-qPCR)^1,4^ or droplet digital polymerase chain reaction (ddPCR)^6,7^. Soon thereafter, genomic surveillance of SARS-CoV-2 was also applied to screen for the presence of SARS-CoV-2 and its variants^8–11^.

Successful SARS-CoV-2 whole genome sequencing from wastewater has been reported sporadically^8–12^, where SARS-CoV-2 reads were successfully mapped to the reference genome at near-full-genome breadth of coverage in considerable read depth. Recently, wastewater SARS-CoV-2 sequencing has also focused on targeted region(s), such as the S gene that contains key mutations for viral evolution and lineage identification^13^. On the other side, mutations in non-spike regions also provide important information about viral replication and transmission^14^. This indicates the importance of whole genome sequencing of SARS-CoV-2 in wastewater samples, which could reveal multiple mutations, variants and lineages circulating in the local communities, and could also be viewed as complementary evidence for novel or emerging variants and lineages in addition to clinical sample sequencing results reported to public health departments^15,16^. Despite the reported successful cases in whole genome and targeted sequencing in the field, sequencing SARS-CoV-2 from wastewater is still very complicated due to significant challenges specific to the sample type^15,17^. These challenges include low target concentration, degraded and/or fragmented RNA, potential PCR inhibitors, and extremely complex microbial and chemical background, which make RNA yield and quality not ideal for downstream sequencing. Sample intrinsic features such as fragmented RNA template are unavoidable. In fact, reports of fragmented wastewater SARS-CoV-2 sequencing results from whole genome amplification are not uncommon^9,18,19^; for example, Amman et al. applied a criteria of 40% genome breadth of coverage for a wastewater sample to be “passed” for downstream sequencing analysis^18^.

To date, there is no standard method for wastewater SARS-CoV-2 sequencing approaches. Various methods can and have been applied for each single step from sample concentration to sequencing, such as concentration methods based on the solids and/or the liquid portion of wastewater^7,20^, extraction methods using a wide variety of commercially available kits with the applications of silica columns or magnetic beads^21^, and various library preparation kits with different sequencing primer schemes (e.g., ARTIC primer schemes^22,23^, QIAseq DIRECT kit^24^, Swift Normalase Amplicon SARS-CoV-2 Panel/IDT xGen SARS-CoV-2 Panel^11,12^), as well as variations in sequencing platforms such as Illumina^11,12^ and Oxford Nanopore Technologies^23^. Whether one or several of these strategies contribute specifically to the reported success for wastewater SARS-CoV-2 genome sequencing is unknown. Here we explored potential causes of incomplete sequencing outcomes in our local wastewater treatment plant (WWTP) influent samples from a technical perspective to improve wastewater sequencing outcomes. We report and compare our sequencing results (i.e., genome breadth of coverage) from WWTP influent composite samples and grab samples in the Chicago area that were collected from March to October 2021, with another two samples from November 2020. In total, three sample processing methods (i.e., concentration and homogenization methods) and two sequencing library preparation kits were tested, yielding mostly incomplete and variable genome breadth of converge using different sample processing and sequencing methods. To explore the potential causes underlying the incompletion and variation of genome breadth of coverage, we examined 1) sequencing outcomes composite and grab samples using different library preparation kits, 2) potential impacts from certain sample intrinsic features, including viral concentration (measured by the CDC N1 assay), flow rate, nutrient concentrations (indicated by the concentration of ammonia nitrogen), biochemical oxygen demand (BOD), and suspended solids content, and 3) the potential contribution of sample processing methods to the sequencing outcomes. We also report and provide a possible explanation for the inconsistency observed between qPCR viral concentrations and sequencing outcomes through the results of a synthetic SAR-CoV-2 RNA control experiment, where different processing methods were tested with raw samples or water (as control) with and without spike-in of full-length synthetic SAR-CoV-2 RNA (Twist Synthetic SARS-CoV-2 RNA Control).

## Material and methods

### Sample collection and processing

Weekly 24-hour raw composite WWTP influent samples were sent on ice to the School of Public Health, University of Illinois Chicago within 4 hours of collection and kept at 4°C before processing. A total of 182 wastewater samples were collected from the Chicago area between November 2020 to October 2021, with most samples collected between March and October 2021, including 99 raw WWTP composite influent and 83 grab samples.

Two concentration methods were used, including the HA filtration method and the Nanotrap method. Samples undergoing HA filtration were processed within 4 hours of arrival. Those undergoing Nanotrap were processed within 12 hours of arrival, prior to which they were stored at 4°C. All sample processing procedures were performed in a biosafety cabinet in a BSL2 laboratory. HA filtration was performed by filtering samples through 0.8 μm cellulose ester HA filters (47 mm diameter; MF-Millipore, Carrigtwohill, Ireland). For Nanotrap, 10 mL of raw sewage was mixed with 150 µL of Nanotrap nanobeads (Ceres Nanosciences, Inc., Manassas, VA) in a 50 mL conical tube and mixed by vortexing briefly. The mixture was then incubated at room temperature for 30 minutes and transferred to a magnetic rack for another 30 minutes until all nanoparticles were aggregated and settled to the bottom where the magnet was positioned. The supernatant was decanted, and the Nanotrap pellets were then eluted with 650 µL viral lysis buffer (Solution PM1 in AllPrep PowerViral DNA/RNA Kit, QIAGEN, Germantown, MD) for downstream extraction.

For HA filter homogenization prior to extraction, bead-beating was employed for viral lysis. We tested two types of beads: the ZR BashingBead Lysis tube (Zymo beads; Zymo, Irvine, CA) and the GeneRite pre-loaded beads tube (glass beads; GeneRite, #S0205-50, North Brunswick, NJ). For both beads, bead beating was performed using a mini beadbeater (Biospec Products, Bartlesville, OK) in two runs of 2.5 min with a 5 min rest in 4°C^6^. Detailed information of samples and processing methods is provided in Supplemental Dataset 1.

### Nucleic acid extraction

Three extraction kits were used: (1) AllPrep PowerViral DNA/RNA Kit (herein “PV”; Qiagen, Hilden, Germany), (2) QIAamp Viral RNA Mini Kit (“QIAamp”; Qiagen, Hilden, Germany), and (3) MegMax Viral/Pathogen Total Nucleic Acid Isolation Kit (“MegMax”; Thermo Fisher Scientific, Inc., Waltham, MA, USA). For HA filtration with Zymo beads (HA+Zymo), the PV kit was used ^6^. For HA filtration with glass beads (HA+glass), the QIAamp Kit was used. For the Nanotrap method, the MegMax Kit was used. All extractions followed the manufacturer’s standard instructions.

Under the same bead beating setting, the Zymo beads homogenized the filter completely to a paste-like texture, while the glass beads left large pieces still intact. Therefore, we consider the HA+Zymo a harsher homogenization method than the HA+glass or Nanotrap method when no beads were used. Also, to interpret impacts from the processing methods’ features, we considered both the PV and QIAamp extraction kits as “silica column-based” extraction methods in our downstream variable of importance selection analysis, and the MegMax kit as a “magnetic beads-based” extraction method. Herein, we refer to the sample “processing methods” as concentration and homogenization methods, and the sample processing groups to “HA+Zymo”, “HA+glass” and “Nanotrap”.

### Synthetic SAR-CoV-2 RNA control testing

In addition to processing real wastewater samples, we also employed controlled testing with synthetic SARS-CoV-2 RNA to elucidate the influence of sample processing methods on sequencing outcomes. We hypothesized that more intensive processing methods (i.e., homogenizing) could have worse recoveries of the free synthetic SARS-CoV-2 RNA in sequencing, and that the wastewater sample context could also affect genome recoveries under the setting of different processing methods.

To test this hypothesis, we spiked 10 µL of 1:10 v/v diluted Twist Synthetic SARS-CoV-2 RNA Control 2 (Twist Bioscience, South San Francisco, CA) into WWTP composite influent samples which then underwent each sample processing combination. The Twist RNA’s concentration was quantified using the N1 assay by testing 1:10 and 1:100 v/v dilutions in duplicate, and was determined to be 1.35 × 10^7^ cp/µL. Four WWTP composite influent samples were used, including two from the Stickney Water Reclamation Plant and two from the Terrence J. O’Brien Water Reclamation Plant, sampled on November 2 and November 9, 2021, respectively. Samples were processed in duplicate using the HA+Zymo, HA+glass, and Nanotrap methods with and without the Twist RNA spiked in. The two November 9 samples using Twist spike-in Nanotrap method were processed in singletons. For controls, Twist RNA was spiked into the same volume of molecular grade water as samples in each processing method. Briefly, two types of positive controls were made for the HA methods in order to evaluate the impacts of filtration on Twist RNA recovery: Twist RNA spiked into 25 mL molecular grade water followed by HA filtration before bead beating (referred as “positive control filtered”), and Twist RNA placed directly in the bead beating tube with a blank filter followed by bead beating (referred as “positive controls not filtered”). For Nanotrap positive controls, Twist RNA was mixed with 10 mL of molecular grade water and was processed as the wastewater samples. All positive controls were in duplicate except for the “positive controls not filtered” that was in singleton due to the limited availability of spiking in material. These method groups resulted a total of 53 extractions for the experiment. Detailed extracts information is listed in Table S1.

### Quantification of SARS-CoV-2 concentration

Quantification of SARS-CoV-2 concentration was performed using quantitative reverse transcription PCR (RT-qPCR) using the CDC N1 assay according to the CDC 2019-nCoV Real-Time RT-PCR Diagnostic Panel ^25^. Briefly, a total reaction volume of 20 µL was used, including 5 µL of the TaqPath™ 1-Step RT-qPCR Master Mix, CG (Thermo Fisher Scientific, Inc.), 1 µL of primers with a final concentration of 500 nM each, and 1 µL of probe with a final concentration of 125 nM, 8 µL DNase/RNase-free water and 5 µL of template. The amplification program started at 25°C for 2 min, followed by 50°C for 15 min and 45 cycles of 95°C for 2 min, then 45 cycles of 95°C for 3 s and 55°C for 30 s. A standard curve was established by running five dilutions of transcribed and purified plasmid DNA targets (Integrated DNA Technologies, Inc, Coralville, IA, USA) in triplicate from 5 × 10^5^ copies to 50 copies per reaction. The N1 assay had a slope of -3.189, y-interception of 41.805, R^2^ of 0.999 and an efficiency of 105.9%.

### Library preparation and sequencing

Two library preparation methods and sequencing platforms were used in this study, including i) the QIAseq DIRECT SARS-CoV-2 Kit (Qiagen, Hilden, Germany) on an Illumina Miseq or Hiseq platform in the Environmental Sample Preparation and Sequencing Facility at Argonne National Laboratory, and ii) the Illumina COVIDSeq Test (RUO Version; Illumina Inc, USA) on an Illumina Nextseq 550 platform performed at the Illinois Department of Public Health (IDPH).

### Metadata collection

Monthly plant operating data for WWTP composite samples metadata analysis were collected by each WWTP and accessed from https://apps.mwrd.org/plant_data/OperatingData.aspx. Metadata collected included air temperature (°F), flow rate (million gallons per day, MGD), suspended solids content (mg/L), biochemical oxygen demand (BOD, mg/L), and ammonia nitrogen (mg/L) on both the sampling day and the day before sampling. For metadata analysis of all 24-hour composite samples, geometric means of two days’ values were used.

### Sequencing data analysis

Raw data were assessed for reads quality in FastQC (https://www.bioinformatics.babraham.ac.uk/projects/fastqc/), followed by adaptor trimming in cutadapt^26^ and quality trimming in BBduk (http://jgi.doe.gov/data-and-tools/bb-tools/). Bwa-mem^27^ was used for paired-end reads mapping to the reference genome of Wuhan-Hu-1 (NCBI RefSeq accession NC_045512.2). Primer trimming was performed with iVar^28^ using bed files specific to the QIAseq DIRECT kit and the Illumina COVIDseq kit, respectively. The trimmed bam file was then realigned using bwa-mem and deduplicated using GATK4^29^ for statistical evaluation and downstream analysis.

### Statistical analysis

Spearman’s rank correlation coefficients were used as a proxy to indicate the relationship between the metadata and sequencing outcomes. Selection of important variables (i.e., feature selection) was performed using the R package ‘Boruta’^30^, which performs random shuffling of the original features (called shadow features) and trains a random forest classifier on this shuffled dataset to evaluate the importance of original features by comparing them to the shadow features (i.e., lower or higher than the importance of ‘Shadow Max’). All statistical analysis was performed using R version 4.0.5^31^.

### Sequencing data access

All sequencing data used in this manuscript are available via the NCBI Sequence Read Archive (SRA) under BioProject PRJNA873764.

## Results

### Incomplete SARS-CoV-2 genomes were recovered from wastewater samples using three sample processing methods and two sequencing library preparation kits

We sequenced 99 raw WWTP composite influent samples and 83 grab samples collected from the Chicago area from November 2020 to October 2021, all but two were collected from March to October 2021 (Table 1; Supplemental Dataset 1). The composite samples were highly variable, with an average N1 concentration of 1.2 × 10^5^ ± 1.4 × 10^5^ copies per liter (cp/L); the grab samples had an average N1 concentration of 4.4 × 10^5^ ± 2.4 × 10^6^ cp/L. Among these samples, 155 were sequenced using the Illumina COVIDseq kit, 25 were sequenced using the QIAseq DIRECT kit, and two were sequenced using both kits, resulting in a total number of 184 sequenced samples. Grab samples were only sequenced using the Illumina COVIDseq kit.

**Table 1.** SARS-CoV-2 sequencing outcomes summarized by sample processing methods and library preparation kits.

| Processing\ sequencing methods | Composite |  |  |  | Grab |  |
| --- | --- | --- | --- | --- | --- | --- |
|  | QIAseq DIRECT kit |  | Illumina COVIDseq kit |  | Illumina COVIDseq kit |  |
| | Genome Breadth of Coverage (Mean ± SD) | Number of Samples Sequenced | Genome Breadth of Coverage (Mean ± SD) | Number of Samples Sequenced | Genome Breadth of Coverage (Mean ± SD) | Number of Samples Sequenced |
| HA+Zymo | 25.2% ± 18.1% | 13 | 12.7% ± 4.2% | 10 | / | / |
| HA+glass | / | / | 49.7% ± 28.3% | 59 | 43.9% ± 29.4% | 83 |
| Nanotrap | 16.9% ± 14.3% | 14 | 28.9% ± 20.0% | 5 | / | / |

For all 184 sequenced samples, an average of 2 M reads were obtained, but sequencing success was highly variable with an average of 2.0 M ± 0.7 M (mean ± SD) from the Illumina COVIDseq kit samples, and 2 M ± 4 M from the QIAseq DIRECT kit samples. We noticed that the percentage of mapped reads differed by kit, regardless of sample processing methods. For Illumina COVIDseq, an average of 9% ± 14% (mean ± SD) of total reads mapped to the reference genome, compared to an average of 88% ± 10% for QIAseq DIRECT. Despite the much lower reads mapping percentage, for the Illumina COVIDseq samples the average final mapped reads number after primer trimming and deduplication was 58.8 fold higher than the QIAseq DIRECT ones. This suggested a high level of PCR duplication using the QIAseq DIRECT kit, which could be due to the low template concentration in wastewater samples and/or the kit’s primer scheme, where multiple primer pairs are designed to target the same regions.

For wastewater SARS-CoV-2 sequencing outcomes, 23 samples (12.5% of all sequenced samples) yielded genome breadth of coverage > 80%, of which three were > 90%. One sample reached near-full genome breadth of coverage (99.2%). However, the majority of samples had incomplete genome recoveries, e.g., 63.6% (n=117) were below 50% breadth of coverage. Regarding the sequencing kits, for Illumina COVIDseq samples, an average breadth of coverage of 43.9% ± 28.7% (mean ± SD) was observed at an average depth of 549X. Among these, the HA+glass samples had on average the highest breadth of coverage (46.3% ± 29.0%), better than the intensive HA+Zymo method (12.7% ± 4.2%, Welch’s t-test, p-value < 2.2 × 10^−6^) and the Nanotrap method (28.9% ± 20.0%), although the latter was not statistically significant (Welch’s t-test, p-value = 0.124) (Table 1). This sequencing outcome difference suggested the potential impacts from sample processing methods and/or sequencing kits, especially the homogenization methods that is the main difference between the HA+Zymo and HA+glass groups. For the QIAseq DIRECT samples, a breadth of coverage of 20.9% ± 16.5% at an average depth of 9,910X was observed. This higher depth was likely due to preferential amplification of some regions over others across the genome and was correlated with the high PCR duplication observed in samples run with this kit. It should be noted that the overall performance of these two sequencing kits were not comparable as there were no HA+glass method processed samples available for the QIAseq DIRECT kit. However, using both kits, the composite samples processed in HA+Zymo and Nanotrap methods both had lower breadth of coverage (Table 1), indicating that the sample processing methods could contribute more to the sequencing outcomes than the sequencing kits. Detailed sample information and sequencing results are listed in Supplemental Dataset 1.

### SARS-CoV-2 whole genome sequencing outcomes were associated with sample types and processing methods

Observing the potential impacts from sample processing method on SARS-CoV-2 genome recovery, we evaluated additional factors that we hypothesized may impact sequencing outcomes, including sample types (influent or grab) and intrinsic features of wastewater samples.

We observed that our grab samples had better correlation between N1 concentration and breadth of coverage (n=83, Spearman’s rho = 0.697, p-value = 2.43 × 10^−13^) than composite samples (n = 59, Spearman’s rho = 0.119, p-value = 0.367) processed using the same method (i.e., HA+glass, Illumina COVIDseq). These grab samples reached an average breadth of coverage of 43.9% ± 29.4% at an average depth of 727X ± 1,656X. Sixteen percent of the grab samples were over 80% breadth of coverage, among which the three highest concentration samples were > 90%, with one of them reached near-full-genome (>99%) breadth of coverage (Figure 1). Also, 24% of the grab samples showed non-detection in RT-qPCR. For composite samples of the same processing methods, an average breadth of coverage of 49.7% ± 28.3% at an average depth of 433X ± 650X was obtained (Table 1). Seventeen percent of them had >80% breadth of coverage, but none reached >90%. No negative samples were detected in composite samples (Figure 1). Our data indicated that these samples’ sequencing outcomes (breadth of coverage) were not significantly different based on sample types when the SARS-CoV-2 concentration was between 10^4^ to 10^6^ cp/L (N1 assay, Welch’s t-test, p-value = 0.07). High SARS-CoV-2 concentration in grab samples (>10^6^ cp/L, N1 assay) very likely played a role in contributing to the near-full-genome breadth of coverage in these HA+glass samples. Yet this observation was not the same for samples using other processing methods. For example, the HA+Zymo method had on average 3-fold higher N1 concentration than the Nanotrap method and 6-fold higher than the HA+glass method (Figure S1) but had the highest breadth of coverage of only 57.7%. Again, this suggested the potential impact of sample processing methods on sequencing outcomes.

**Figure 1.**
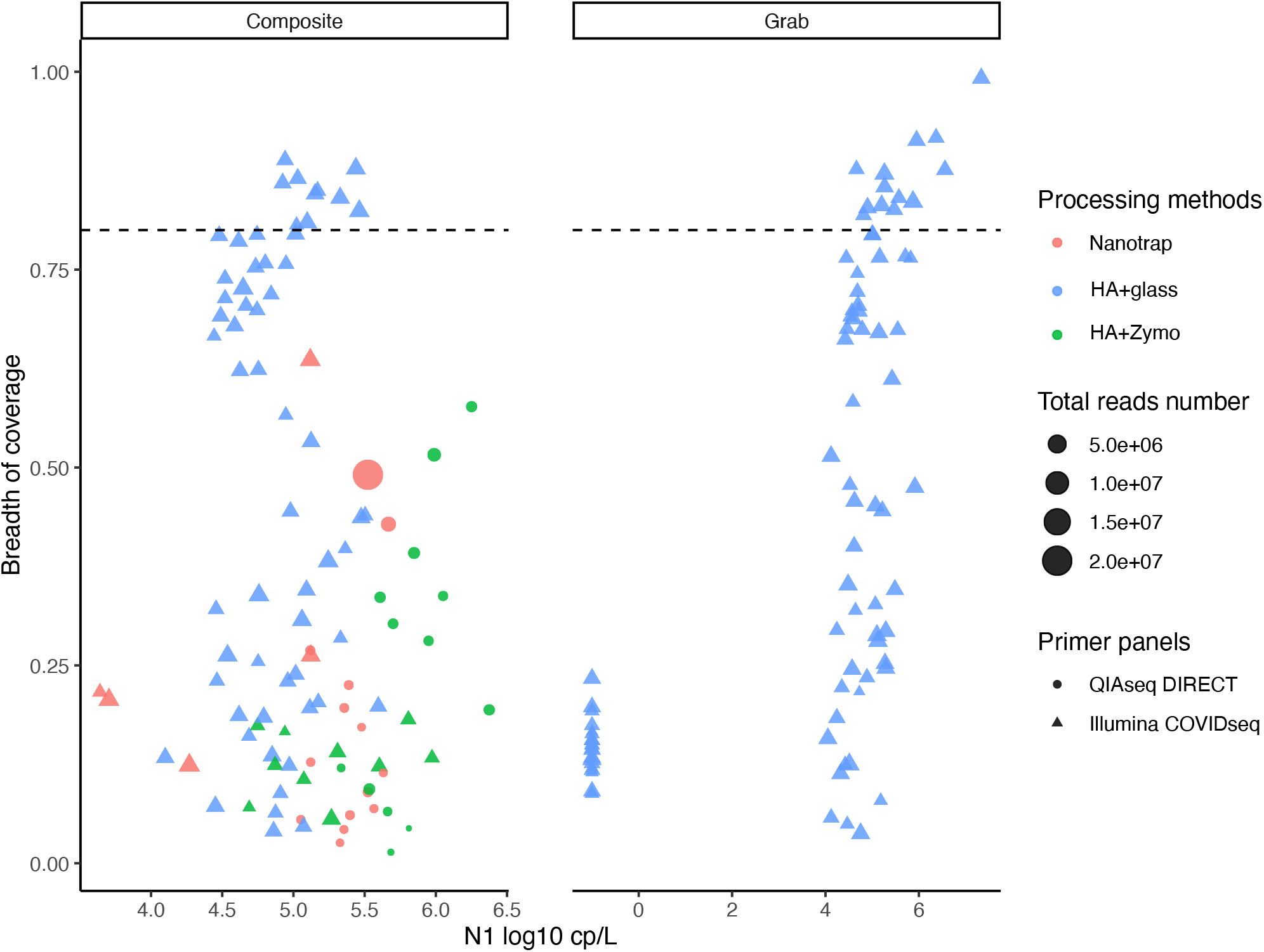
Comparison of N1 gene copy numbers via RT-qPCR and SARS-CoV-2 genome breadth of coverage via sequencing in composite and grab sewage samples, using the QIAseq DIRECT kit and Illumina COVIDseq kits. The x-axis represents the N1 concentration (gene copies) per liter of sewage sample, and the y-axis represents for the breadth of coverage observed for each sequencing run. The dashed line represents a genome breadth of coverage of 80%. The dot colors indicate the sample concentration and homogenization method, the sizes represent the sequencing depth of each sample, and the shapes indicate the library preparation kit used.

We then examined the potential impact on the sequencing outcomes from the wastewater samples’ intrinsic features, including the SARS-CoV-2 concentration (cp/L, N1 assay), air temperature (°F), flow rate (MGD), suspended solids content (mg/L), BOD (mg/L), and ammonia nitrogen (mg/L) (Supplemental Dataset 1). We collected all available metadata for the HA+glass method processed composite samples (n=55) and examined their relationships with samples’ genome breadth of coverage using both the Spearman’s rank correlation test and random-forest-based selection of important features (i.e., Boruta). Spearman’s correlation analysis showed that among all the WWTP reported metadata, only the flow rate had a weak yet not statistically significant correlation with the breadth of coverage (Spearman’s rho = 0.251, p-value = 0.064) (Figure S2A).

Random forest-based feature selection method (i.e., R package ‘Boruta’) was used to further evaluate the potential important variables in samples’ metadata (e.g., flow rate, temperature, BOD, ammonia nitrogen and suspend solids contents) and samples processing methods (processing methods, extraction methods and library preparation kits) for composite samples’ sequencing outcomes. Our results showed that only the processing methods (i.e., concentration and homogenization) and library preparation kits were important features, which exceeded the “Shadow Max” value that represents for the highest importance of the shadow features. The processing methods had much higher importance value than the shadow max value compared to the library prep kit, indicating the important roles of concentration and homogenization in contributing to sequencing outcomes (Figure 2). Interestingly, the extraction method type (i.e., silica column-or magnetic beads-based) was not considered important, again suggested that the input RNA quality was most consequential in impacting sequencing outcomes. In addition, feature selection using sample’s metadata only confirmed no intrinsic feature was considered important (Figure S2B). Overall, this analysis suggested that for composite samples, the processing methods (concentration and homogenizing methods) are likely determining factors for genome breadth of coverage. Library preparation kit could also play a role; extraction method or sample intrinsic features is likely less influential.

**Figure 2.**
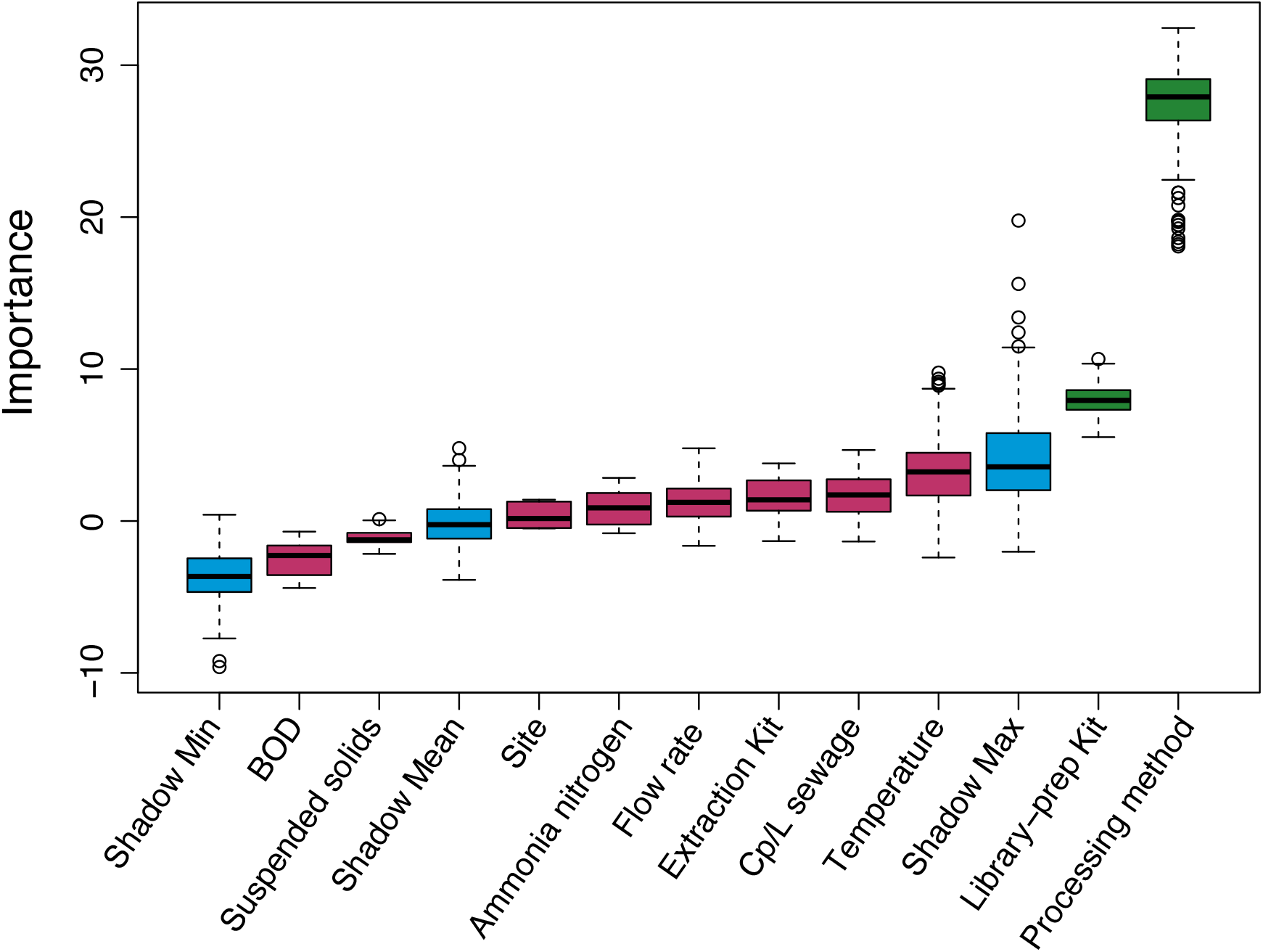
Statistical assessment of impact of sampling and processing variables on sequencing breadth of coverage using all composite samples with available metadata. The “Shadow Min”, “Shadow Mean” and “Shadow Max” values indicate the minimal, average and maximum Z score of a shadow feature decided by Boruta, respectively (Blue boxes). Features with an importance metric that exceeds the Shadow Max value are considered important (green boxes), and features with importance below the Shadow Max value are deemed not important (red boxes). In this analysis only processing method (i.e., concentration and homogenization) and library preparation kit are considered important by Boruta.

### Potential mechanisms for impact of processing methods on sequencing outcomes, elucidated by a synthetic RNA spike-in experiment

To confirm that sequencing outcomes of SARS-CoV-2 in wastewater samples could be impacted by the yielded RNA quality from different processing methods, we conducted a synthetic SARS-CoV-2 RNA test. We spiked about 10^7^ copies of the Twist SARS-CoV-2 synthetic RNA control into composite sewage samples and molecular biology grade water, respectively, and processed using HA+Zymo, HA+glass, and Nanotrap methods. The samples were extracted, quantified, and sequenced for genome breadth of coverage evaluations.

From RT-qPCR results (N1 assay), all Twist-spiked samples had higher concentrations compared to their no spike-in pairs. The Nanotrap group samples spiked with Twist had on average 1.2 folds recovery of the no spike-in group, the HA+glass group samples had on average 5.0 folds, and the HA+Zymo group was on average 13.1 folds of the no spike-in group. This suggested all three sample processing methods recovered Twist RNA to some extent, with the highest capacity being the HA+Zymo method (Figure 3). However, when compared to the spiked-in concentration, all three methods had on average very low recovery of the Twist RNA (HA+Zymo 2.21%, HA+glass 0.17%, Nanotrap 0.05%). The highest recoveries of Twist RNA were the positive controls (filtered) in the HA+glass group, with an average of 19.2% recovered of the original spike-in. The Nanotrap method had overall the least N1 concentration in both the sample groups and the positive controls, indicating the nanoparticle’s limited capability to trap the total nucleic acids in this study.

**Figure 3.**
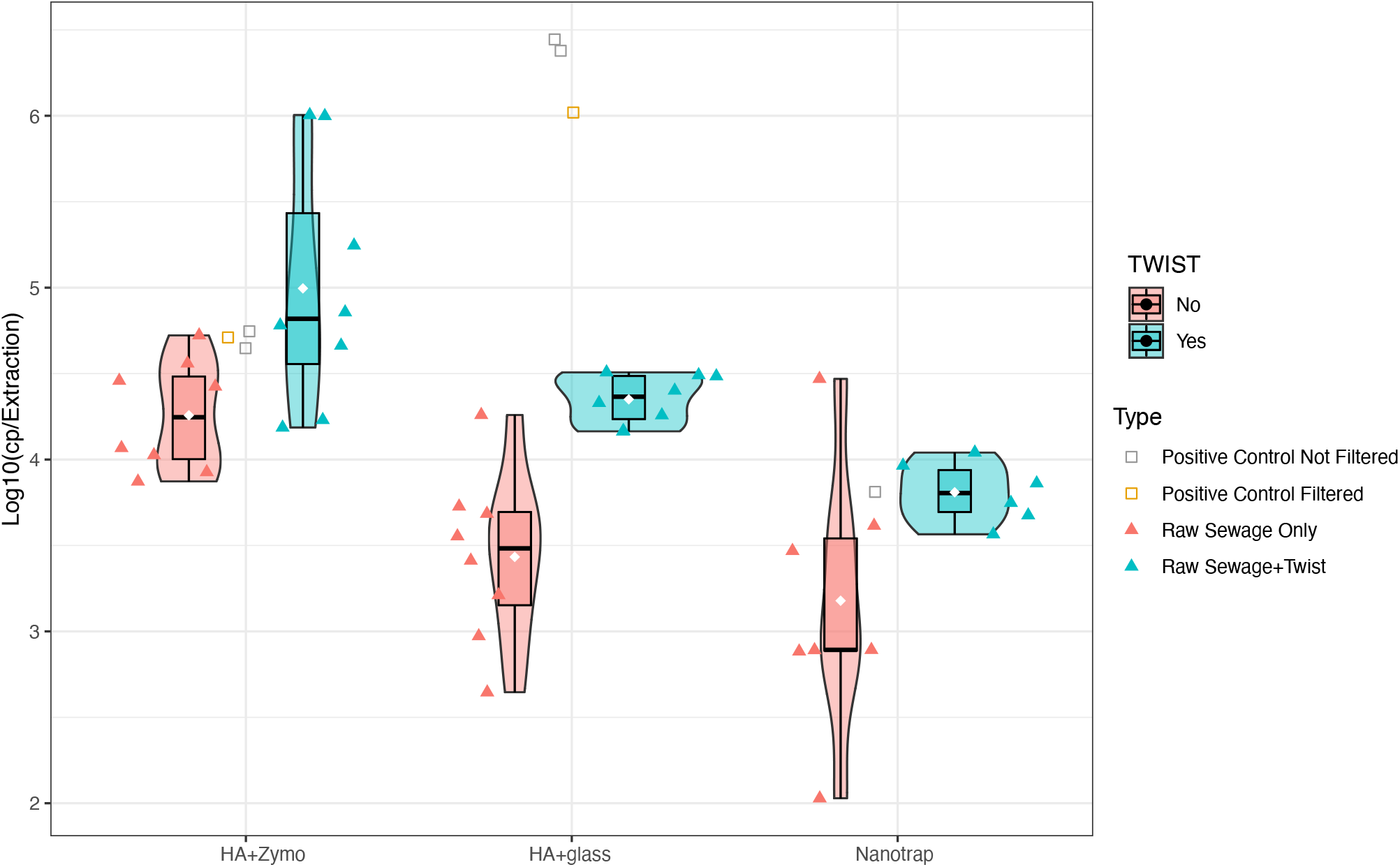
RT-qPCR results of Twist positive control, Twist-spiked-in and no-spiked-in samples. The y-axis shows the SARS-CoV-2 concentration in N1 copies per processed sample volume, or per extraction. Processing methods are indicated on the x-axis. The squares indicate positive controls of Twist spike-in, including positive control processed without filtration and positive control processed with filtration. Triangles represent for samples with or without Twist spike-in. The boxplot whiskers show the 25^th^, 50^th^ and 75^th^ percentile of each group. The white diamond shows the mean value of all data points in each group.

Like our previous observation of N1 concentration in composite samples (Figure S1), the HA+Zymo method had the highest N1 concentration in samples both with and without Twist RNA (Figure 3), indicating its higher capacity of releasing and/or grabbing the total amount of N1 target, likely through the complete filters’ disintegration. Interestingly, although the HA+glass group had lower N1 concentration in samples compared to the HA+Zymo group, it showed 1.7 orders of magnitude higher concentration in the positive controls. This indicated that the two processing methods behaved differently in recovering nucleic acids depends on the sample context, i.e., when from a mixture of sample and free RNA or when just free RNA.

We then sequenced all the extracts from the synthetic RNA test (Figure 4). As expected, the HA+Zymo method positive controls had much less genome breadth of coverage (47.57% ± 14.63%) compared to the HA+glass or Nanotrap methods’ positive controls (both >95%). Also, the mean coverage of the HA+Zymo group positive controls was 549X, while the HA+glass positive controls reached > 15,300X and the Nanotrap group also reached 7,800X. This supported our hypothesis that the HA+Zymo method, which is more harsh in homogenizing the filters, could have RNA templates of worse quality (e.g., more fragmented) compared to the other two groups despite its much higher N1 concentration; likely this could be attributed to the impacted genome integrity. It is also worth noting that the Nanotrap group positive controls also reached > 95% genome breadth of coverage with >10 folds higher depth than the HA+Zymo group positives, despite its much lower N1 concentration (Figure 3). This further suggested that the RNA quality could be the key factor impacting sequencing outcomes.

**Figure 4.**
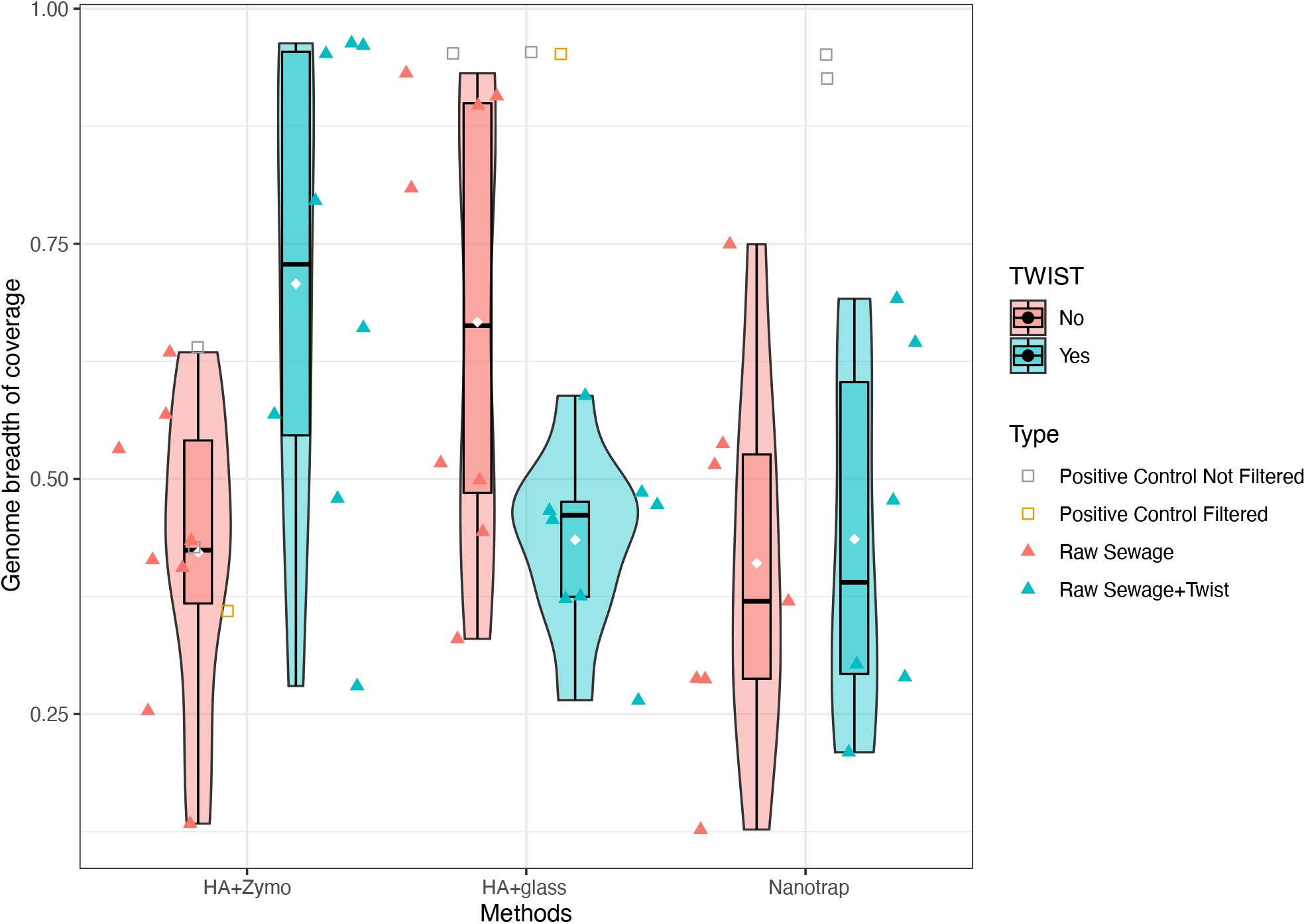
Sequencing results of Twist positive control, no-spiked-in and Twist-spiked-in samples. Y-axis represents the genome covered percentage. X-axis represents processing methods. The squares indicate positive controls of Twist, including positive control (Nanotrap or without filtration for HA) and positive control filtered (with water using HA filtration). Triangles show samples with or without Twist spike-in. The boxplot whiskers represent the 25^th^, 50^th^ and 75^th^ percentile of each group. The white diamond stands for the mean value of all data points in each group.

All three sample groups with Twist spiked-in had overall 1.4 ± 0.3 (mean ± SD) folds higher genome breadth of coverage values than their paired groups without Twist, and 6.5 ± 6.1 folds higher N1 concentration than their paired groups, indicating the spiked-in Twist RNA was recovered to some extent by all methods. The Nanotrap group had the lowest genome percentage recovered in samples compared to the other two, which was consistent with the qPCR results and could be due to the limited capacity of nucleic acid adsorption of this method. The HA+glass and HA+Zymo groups showed overall similar mean genome breadth of coverage in sewage samples with Twist (mean value 65.7% and 70.8%, respectively) from the two biological samples. However, considering that the HA+glass group had on average 12 times less N1 concentration than the HA+Zymo group (Figure 3 and 4), this indicated that the less intensive sample processing method could contribute to a similar genome recovery level at a much lower template concentration (indicated by RT-qPCR), and again, suggested the inconsistency of qPCR quantification and sequencing genome recovery. In addition, both the HA+glass and Nanotrap sample groups had lower genome breadth of coverage values compared to their positive controls, indicating that the sample matrix could have adverse effects in recovering total RNA compared to when there was just free RNA in these methods. In contrast, in the HA+Zymo group, the raw sewage+Twist treatment had 1.5 folds higher average genome breadth of coverage than the positive control samples, and 1.7 folds higher than the raw sewage only samples, indicating that the sample matrix could contribute to preserving the spiked-in free RNA in this more intensive method. More studies are needed to better understand how wastewater matrix could affect the quality of extracted RNA under different processing methods.

## Discussion

Wastewater epidemiology (WBE) has been demonstrated to be a useful and successful tool for surveillance of SARS-CoV-2 genetic signals in a community-wide scope^2,3,32^. The change in SARS-CoV-2 genetic signals overtime could show the overall local trend of COVID-19 infection, providing complementary public health information to clinical diagnostic tests^2,3,32^. The most widely adopted WBE as of today is RT-qPCR/ddPCR/dPCR where the level of detected SARS-CoV-2 could be reported^32,33^. Further, with the applications of variant-specific assays, the quantification of a variant’s presence has also been realized^34^. While quantification-based methods provide useful information for existing SARS-CoV-2 variants that are already known to be circulating in a community, whole genome sequencing of SARS-CoV-2 from wastewater is attractive because it not only provides higher resolution of the known circulating variants, but also enables broad identification of novel mutations within the context of the whole genome, therefore contributes to understanding the viral evolution and/or transmission within the sewersheds. However, sequencing SARS-CoV-2 from wastewater matrix is challenging due to issues such as low RNA concentration, degraded RNA, and complex microbial background^15,17^.

Our study provides evidence that the wastewater sample processing methods, including concentration and homogenization procedures, could lead to an inconsistency in RT-qPCR measurement and sequencing outcomes. We picked the methods of HA filtration, which requires bead beating, and Nanotrap that has no intensive sample mixing, as a means to explore the impacts of the intensity of processing methods on sequencing results. The wastewater samples’ SARS-CoV-2 N1 gene concentrations identified by qPCR were positively correlated with the intensity of processing method (Figure S1, Figure 3), indicating that the HA+Zymo method released more N1 gene than the HA+glass and Nanotrap methods. However, the same processing method did not outperform, or was even worse than, the other two in sequencing outcomes (breadth of coverage) (Figure 1, Figure 4). This suggested an inconsistency between the RT-qPCR measurement of a single gene and the sequencing outcomes, which requires the amplification across the whole genome. Interestingly, we did observe that our samples’ SARS-CoV-2 concentrations (as measured by RT-qPCR) associated with a higher genome breadth of coverage only using the HA+glass method; for example, the >90% breadth of coverage samples all had the concentrations > 10^6^ cp/L (Figure 1). Based on this result, it is likely that the RNA templates released by the intensive processing method (HA+Zymo) may not be suitable for sequencing, which should have amplicons of ∼400 bp, and may have caused the incomplete genome breadth of coverage. It is reasonable that the HA+glass method could have released RNA template of better quality, as the bead beating intensity along with the produced heat in the HA+Zymo method could lead to more fragmented RNA templates than the HA+glass and Nanotrap methods, which are of lower homogenizing intensities. This observed inconsistency of RT-qPCR and sequencing outcomes using the intensive method also indicated that selection of wastewater samples for sequencing should not solely depend on concentration quantified by RT-qPCR, as not enough information about the overall RNA quality is indicated. To maximize the efficacy of wastewater SARS-CoV-2 quantitative and genomic surveillance, a suitable sample processing method that balances the need for sufficient RNA template yield while not negatively impacting the obtained RNA quality (over-fragmentation) is essential. For example, automatic RNA extraction was reported to have better recovery for RNA extraction from the human influenza virus and respiratory syncytial virus compared to manual extraction^35^, and has been adopted by a successful wastewater genome surveillance study (e.g., KingFisher Flex system)^12^ as well as other quantitative surveillance studies ^21,36^. Interestingly, in our study, the RNA extraction kits were thought to not impact the sequencing outcomes (via Boruta’s feature selection); this agrees with Qiu et al.’s observation, where the authors have compared five commercial RNA isolation kits, including the three used in our study (QIAamp, MagMax, and PowerViral that shares the same buffers as the PowerMicrobiome kit tested by Qiu et al.), that these kits showed comparable recovery rate for their surrogate human coronavirus 229E^37^.

Our comparison of composite and grab samples sequencing results suggested that with a less intensive processing method (HA+glass), the samples type could affect the sequencing outcomes. It is worth noting that most samples >80% breadth of coverage had a target concentration of >10^5^ cp/L in both composite and grab samples. However, only the grab samples had near-full genome breadth of coverage (over 90%, n=3; Figure 1) or low coverage with no RT-qPCR detection (n=20). Therefore, the difference in sample types (composite and grab sample) could also impact SARS-CoV-2 whole genome sequencing outcomes in at least some cases.

We also explored the impacts of intrinsic wastewater sample features on sequencing outcomes (e.g., flow rate, solids content, chemicals, temperature, nutrients). Our results from both the random forest-based feature selection analysis and the Spearman’s rank correlation analysis suggest that wastewater parameters monitored over the course of this study were mostly not significantly correlated to sequencing outcomes. This result agrees with Kevill et al.’s finding that turbidity and temperature of wastewater samples are not significant factors affecting SARS-CoV-2 genome recovery^38^. However, the flow rate showed a weak positive correlation with the sequencing breadth of coverage using Spearman’s rank correlation analysis. Flow rate is generally negatively correlated with residence time^39^, therefore it is possible a higher flow rate is correlated with better SARS-CoV-2 genome recovery in sequencing. In addition, other environmental factors in the sewer environment, such as sewer biofilm that has been reported to contribute to the decay of human coronavirus^40^, could also contribute to more RNA degradation when the residence time increases. The shorter residence time of the grab samples (<24 hours) compared to the composite samples (1-3 days) could contribute to our near-whole-genome sequencing results in the grab high concentration samples in this study as well. Overall, these observations support the idea that longer residence time could cause decayed RNA in the wastewater, impacting the downstream RNA extraction and amplicon sequencing. Additional studies are needed from different geographical regions to study how environmental factors in sewers affect SARS-CoV-2 genome sequencing in wastewater samples.

## Conclusions

In this study, we explored the impacts from sample types, sample intrinsic features and most importantly, sample processing methods on wastewater SARS-CoV-2 amplicon sequencing outcomes. We identified that the sample concentration and homogenization methods play major roles impacting downstream incomplete genome recoveries and could contribute to the inconsistency of RT-qPCR concentrations and sequencing outcomes. More studies of different techniques and wastewater in other geographical regions are needed for standardizing successful whole genome amplicon sequencing in the field.

## Data Availability

All data produced in the present work are contained in the manuscript.

## Acknowledgement

This study was funded by the Walder foundation. We thank Dr. Ira Heimler at the Illinois Department of Public Health (IDPH) for performing timely sequencing work using the Illumina COVIDseq kit for this study.

