## Supplemental figures and table for "Intensity of sample processing methods impacts wastewater SARS-CoV-2 whole genome amplicon sequencing outcomes"

**List of supplemental figures and tables**

Figure S1. N1 concentrations of all samples processed using three processing methods.

Figure S2. Analysis of impacts from the wastewater sample intrinsic features on sequencing outcomes.

Table S1. Samples and settings for the synthetic SAR-CoV-2 RNA control testing.

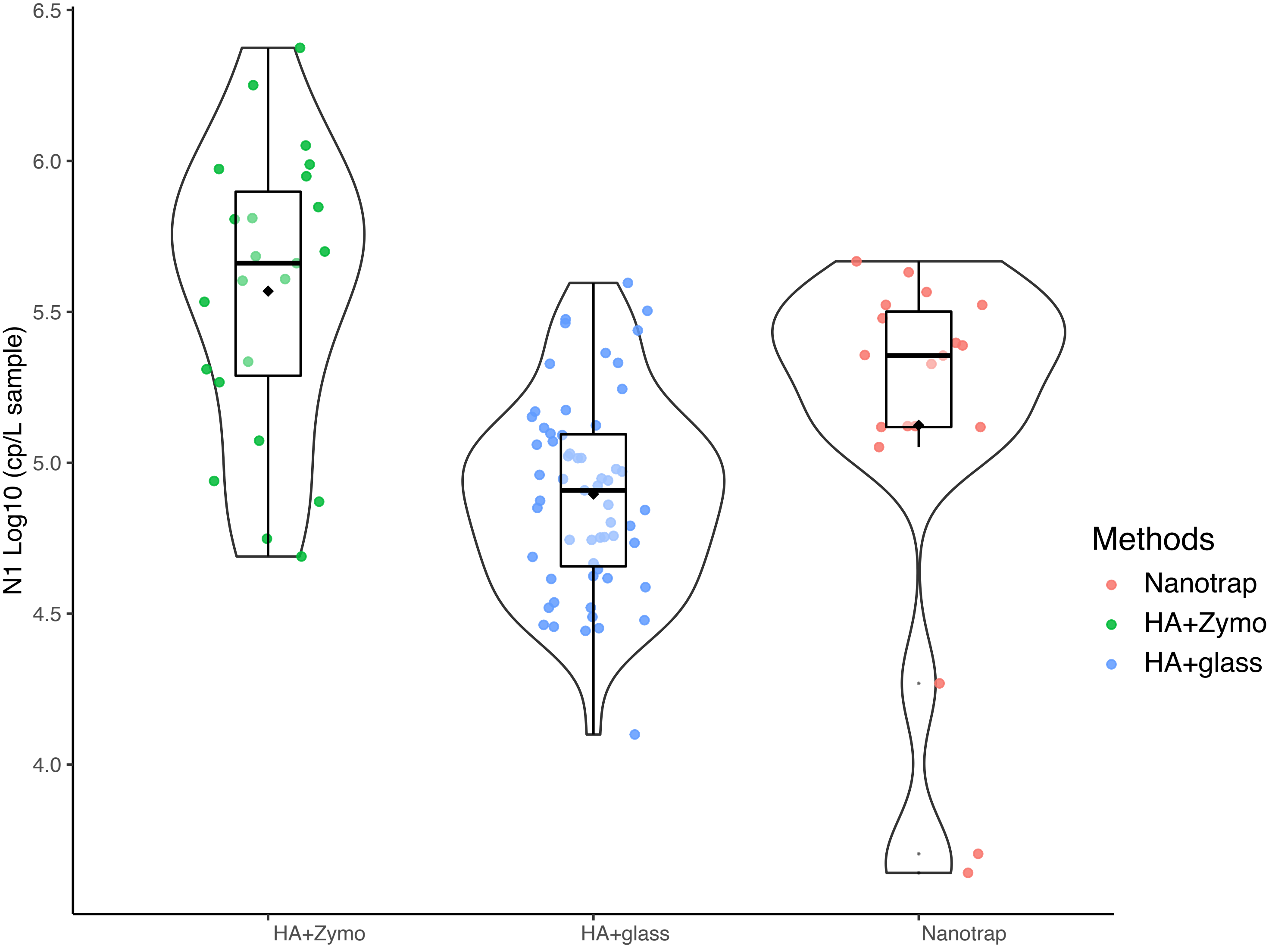


Figure S1. N1 concentrations of all samples processed using three methods. X-axis shows the processing methods used, y-axis shows log10 transformed N1 concentration in copies per liter samples. The boxplot whiskers show the 25$^{th}$, 50$^{th}$ and 75$^{th}$ percentile of each group.

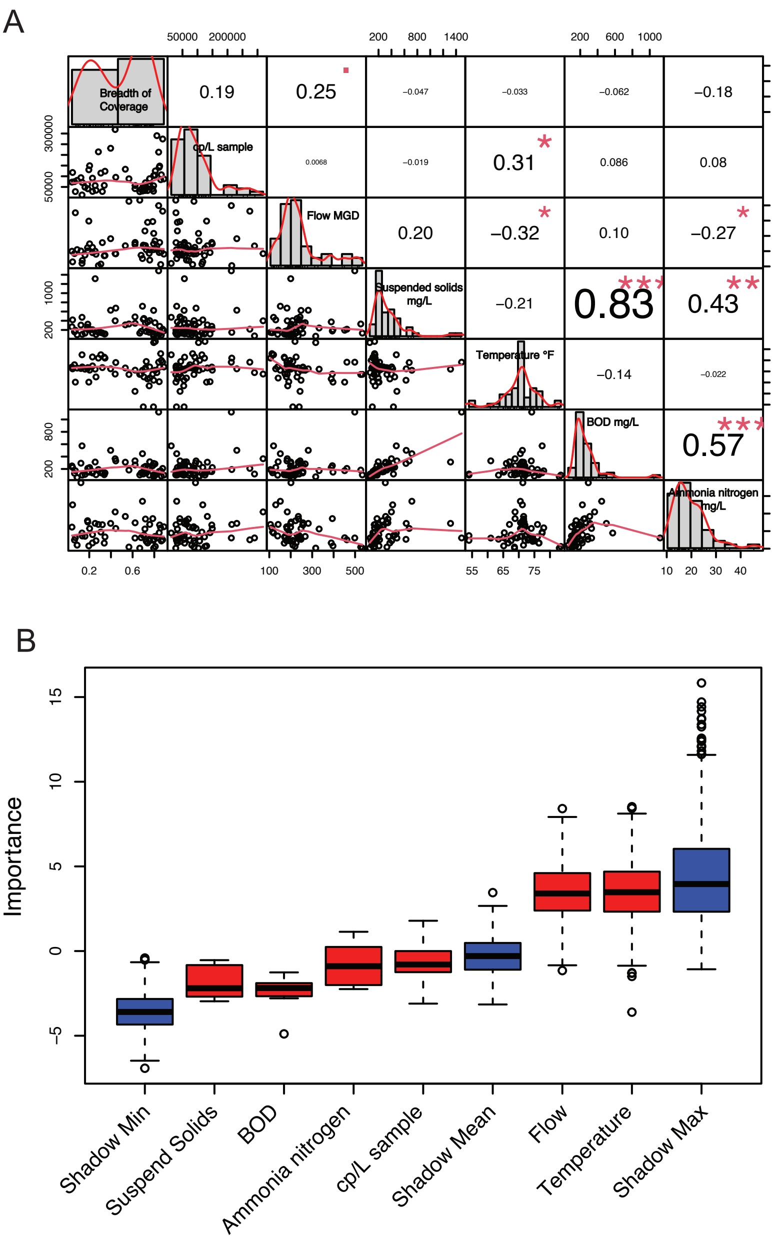


Figure S2. Analysis of impacts from the wastewater sample intrinsic features on sequencing outcomes. A). Spearman's correlation analysis indicates only the flow amount has weak correlation with sequencing breadth of coverage. B). Boruta's feature selection analysis indicates no intrinsic feature is important for impacting sequencing outcomes.

**Table S1. Samples and settings for the synthetic SAR-CoV-2 RNA control testing.**

| Sampling site | Sampling Date | Twist | Method | Extracts number |
|---|---|---|---|---|
| Terrence J. O'Brien | 11/2/21 | No | HA_Glass | 2 |
| Terrence J. O'Brien | 11/2/21 | No | HA_Zymo | 2 |
| Terrence J. O'Brien | 11/2/21 | No | NT | 1 |
| Terrence J. O'Brien | 11/2/21 | Yes | HA_Glass | 2 |
| Terrence J. O'Brien | 11/2/21 | Yes | HA_Zymo | 2 |
| Terrence J. O'Brien | 11/2/21 | Yes | NT | 2 |
| Terrence J. O'Brien | 11/9/21 | No | HA_Glass | 2 |
| Terrence J. O'Brien | 11/9/21 | No | HA_Zymo | 2 |
| Terrence J. O'Brien | 11/9/21 | No | NT | 2 |
| Terrence J. O'Brien | 11/9/21 | Yes | HA_Glass | 2 |
| Terrence J. O'Brien | 11/9/21 | Yes | HA_Zymo | 2 |
| Terrence J. O'Brien | 11/9/21 | Yes | NT | 1 |
| Stickney | 11/2/21 | No | HA_Glass | 2 |
| Stickney | 11/2/21 | No | HA_Zymo | 2 |
| Stickney | 11/2/21 | No | NT | 2 |
| Stickney | 11/2/21 | Yes | HA_Glass | 2 |
| Stickney | 11/2/21 | Yes | HA_Zymo | 2 |
| Stickney | 11/2/21 | Yes | NT | 2 |
| Stickney | 11/9/21 | No | HA_Glass | 2 |
| Stickney | 11/9/21 | No | HA_Zymo | 2 |
| Stickney | 11/9/21 | No | NT | 2 |
| Stickney | 11/9/21 | Yes | HA_Glass | 2 |
| Stickney | 11/9/21 | Yes | HA_Zymo | 2 |
| Stickney | 11/9/21 | Yes | NT | 1 |
| positive control filtere | / | Yes | HA_Glass | 1 |
| positive control filtere | / | Yes | HA_Zymo | 1 |
| positive control not filt | / | Yes | HA_Glass | 2 |
| positive control not filt | / | Yes | HA_Zymo | 2 |
| positive control | / | Yes | NT | 2 |
